# Epidemiology of Congenital Heart Disease in Qatar: Preliminary findings from the Sidra Medicine Cardiac Registry

**DOI:** 10.1101/2023.08.24.23294593

**Authors:** Muna Almasri, Khalifa Al-Sulaiti, Dana Al Sayegh, Omna Sharma, Aya El Jerbi, Zakariya Al-Riyami, Padma Kumari Sarada, Samir Gupta, Hesham Al-Saloos, Mange Manyama, Kholoud N. Al-Shafai

**Author notes:** **Corresponding author:** Kholoud N. Al-Shafai, PhD, Translational Medicine Division, Sidra Research, Sidra Medicine., P.O. Box 26999, Doha, Qatar.

## Abstract

**Background:** Congenital heart disease (CHD) is one of the most common congenital anomalies in newborns. The birth prevalence of CHD across the world is variable, reflecting differences in diagnostic capabilities as well as environmental and/or genetic risk factors between regions. Here, we aim to determine the epidemiology of CHD in Qatar in terms of the magnitude, subtypes, associated risk factors and complications.

**Methods and results:** Data for 76 subjects with CHD were extracted from the Sidra cardiac registry database in October 2022. Extracted information for each subject included on age, nationality, gender, CHD diagnosis, outcomes of CHD, and comorbidities. Maternal history for some of the risk factors were also obtained from the registry. Statistical analysis was performed using Chi-Square tests and Fisher’s Exact tests.

Most of the subjects were from the Middle East and North Africa (MENA) region (52.6%), followed by Southeast Asian (30.3%) and Qatari (14.5%). The three most common CHD types were ventricular septal defect (VSD), Atrial septal defect (ASD), and Transposition of the great arteries (TGA). About 32% subjects were born by parents who are first-degree cousins. This association was statistically significant (P<0.05). About 6.9% subjects in this cohort were born to mothers who had Pre-Gestational Diabetes Mellitus.

**Conclusion:** The most common CHD type in this study was VSD followed by ASD. The rate of parental consanguinity was generally high among patients with CHDs. The pattern of non-genetic risk factors, comorbidities, and outcomes in patients with CHD was similar to those previously reported in other studies.

**Clinical perspective:**

- Congenital heart diseases and other birth defects are largely understudied in Qatar. This study provides epidemiological characteristics of patients with congenital heart diseases in Qatar.
- A significant number of subjects in this study had multiple CHD lesions.
- This study highlights the association between parental consanguinity and the increased incidence of congenital heart diseases in Qatar, a country with a high rate of parental consanguinity.
- Majority of the subjects in this study underwent surgical intervention, perhaps reflecting the complexity of the CHD lesions.

## Introduction

The incidence rate of congenital heart disease (CHD) is estimated to be 17.9/1000 worldwide, with ventricular septal defect and atrial septal defect being the most common subtypes accounting for about 29.6% of all cases of CHD.^1^ Asia ranks as the region with the highest cases of congenital heart defects (9.3 per 1000 live births), followed by Europe (8.2 per 1000 live births) and lowest rate is in Africa (1.0 per 1000 live births).^2^ Since 2010, there has been an approximately 5% increase in CHD prevalence every five years, which is likely due to increased detection and diagnosis of milder CHD lesions, as well as greater usage of echocardiography with improved technique.^2^

CHDs can have a major impact on the well-being of the newborn and the parents. In the United States of America, congenital heart diseases are responsible for the largest proportion of up to 50% of mortality caused by birth defects among infants and young children.^3,4^ Before cardiac surgery, 30% of children with severe CHD did not survive until adulthood.^5^ Countries that have structured their care network following this evolution design have substantially increased the survival of children with severe CHD. Despite the progress, CHD is still associated with 85% of death in stillbirths, newborns, and infants, with cardiac arrest from arrhythmias as the main cause.^6^ Patients with CHD may also present with cardiac or non-cardiac comorbidities, which may complicate the management care in this population.

The etiology of congenital heart defects is thought to be multifactorial, with both genetic and environmental risk factors being implicated.^7^ While there is no clear inheritance pattern owing to the different types of CHD and the various environmental factors that predispose to CHD, there is likely two-hit pathogenesis wherein the genetic pre-disposition and additional non-genetic risk factors such as maternal diabetes and obesity can put the fetus at a higher risk of developing CHD.^8^ Our previous study on patients with CHD in Qatar identified cytogenetic abnormalities, pathogenic Single-Nucleotide Variants, and some recessive variants, such as c.884A>G in SMYD6 as the potential genetic etiology of CHD.^9^

Several non-genetic and modifiable risk factors play a role in the development of CHD. These include older maternal age, maternal diabetes (including pre-gestational and gestational), hypertension, use of certain medications during pregnancy such as lithium, anti-epileptics, and antibiotics, and alcohol.^10,11^ The modifiable risk factors are more crucial to identify at an early stage to prevent development of CHD in the newborn. Parental consanguinity has also been associated with CHD as a risk factor, especially in regions where endogamy is a common practice.^12^

The outcomes in patients with CHDs vary widely from spontaneous cure to death.^13^ This is partly due to the differences in the severity of CHDs. The most common interventional treatment modalities include surgical (corrective or palliative), and interventional catheterization. About 15% of patients with CHD who undergo corrective surgery, require re-operation.^14^ Cyanotic conditions such as Tetralogy of Fallot (ToF), Dextro-Transposition of the Great Arteries (D-TGA), and single ventricle have great variation in complexity and severity and hold different approaches to surgical intervention.

Published reports on the epidemiology of CHD in Qatar in terms of the magnitude, major subtypes, associated risk factors and long-term complications is limited. This retrospective study was conducted to provide more information on the epidemiological characteristics of congenital heart diseases in Qatar.

## Methods

### Study Subjects

Data for this study was obtained from the Cardiac Registry at Sidra Medicine. Sidra Medicine is a large quaternary care Children’s hospital in Doha, Qatar. The Heart Center at Sidra Medicine provides preventative, medical, and surgical care for patients with heart diseases including children with congenital or acquired heart conditions.

The cardiac registry is an electronic database established in July 2019 for research purposes. It continues to register cardiac patients who meets inclusion criteria. Only patients with CHD in the database were included in this study.

Extracted de-identified information for each patient included age, nationality, gender, CHD specific diagnosis, outcomes of CHD treated within the first six months of life, and comorbidities. Maternal history including age at delivery, maternal diabetes mellitus, alcohol use, and parental consanguinity were also obtained from the registry. Diagnosis of CHD in each patient was either diagnosed prenatally or in infancy, confirmed by an experienced cardiologist utilizing one or more of the following diagnostic tools: echocardiogram, cardiac Magnetic resonance imaging (MRI), computerized tomography (CT) angiography, or diagnostic catheterization. All CHD cases were classified using ICD-10-CM (International Classification of Diseases, 10th Revision, Clinical Modification).

### Classification of Cardiovascular Malformations

Because most of the subjects in this study had multiple CHD lesions, anatomical and severity categorizations were utilized to group patients into two groups (Table 1 and 2). The anatomical classification is based on the presumed embryological timing. This classification is based on anatomical (not clinical or hemodynamic) criteria.^9^ This classification has been utilized by previous studies.^15,16^ In this classification, patients could be included in more than one embryological category depending on the major defects possessed (this is a modification of the practice done by the origin studies, as several patients in our registry had multiple complex CHDs belonging to more than one category). In the severity scale, patients with CHD were classified using a previously defined hierarchical severity ranking based on the perinatal mortality rate for each of the subgroups, ranging from SI (high perinatal mortality) to SIII (low perinatal mortality).^17^ Patients with multiple CHDs, were grouped into one severity category by taking the specific anatomic CHD type with the greatest perinatal mortality into consideration.

**Table 1.**
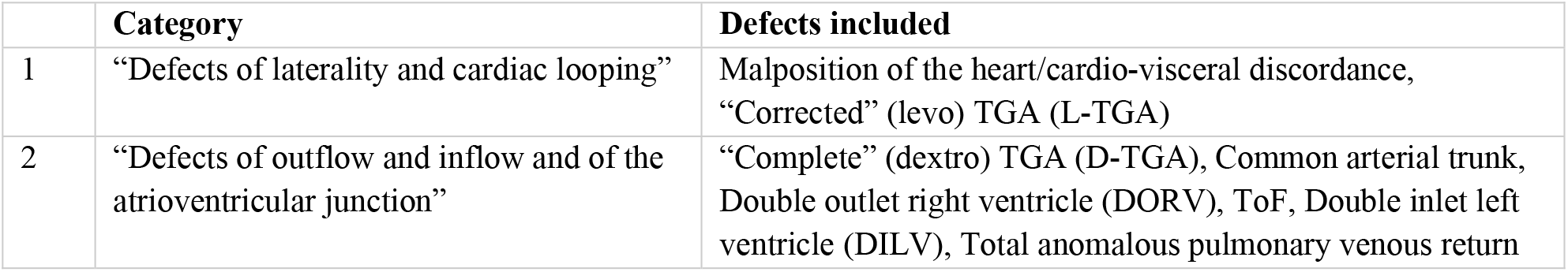
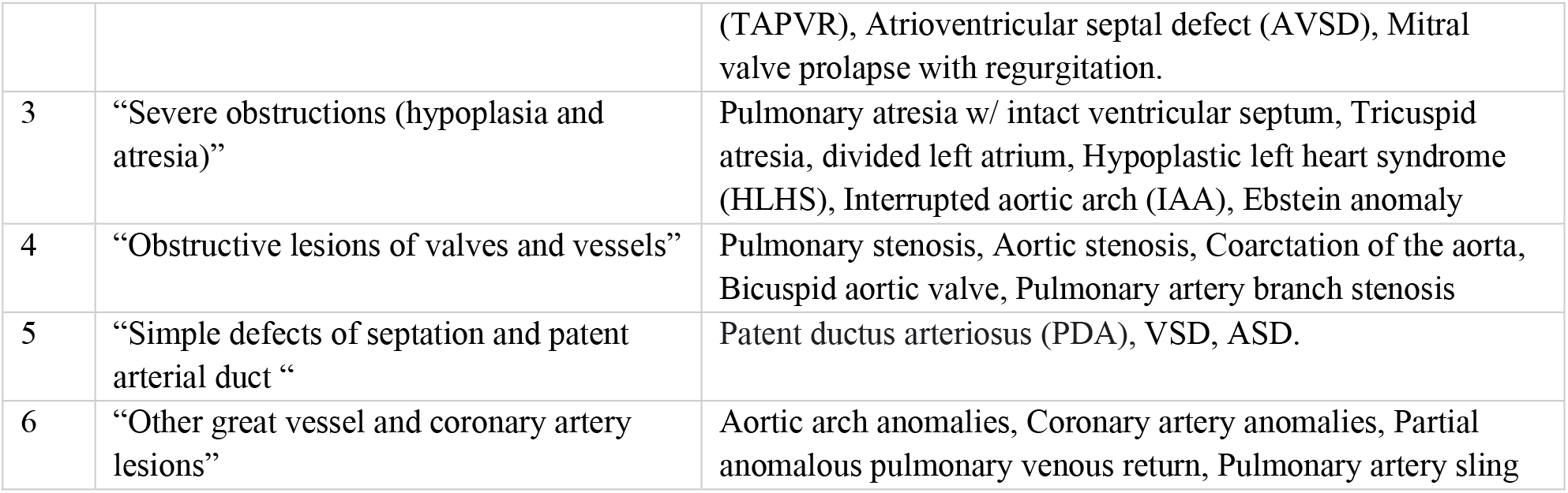
Anatomical categorization.

|  | Category | Defects included |
| --- | --- | --- |
| 1 | "Defects of laterality and cardiac looping" | Malposition of the heart/cardio-visceral discordance, "Corrected" (levo) TGA (L-TGA) |
| 2 | "Defects of outflow and inflow and of the atrioventricular junction" | "Complete" (dextro) TGA (D-TGA), Common arterial trunk, Double outlet right ventricle (DORV), ToF, Double inlet left ventricle (DILV), Total anomalous pulmonary venous return |
|  |  | (TAPVR), Atrioventricular septal defect (AVSD), Mitral valve prolapse with regurgitation. |
| 3 | “Severe obstructions (hypoplasia and atresia)” | Pulmonary atresia w/ intact ventricular septum, Tricuspid atresia, divided left atrium, Hypoplastic left heart syndrome (HLHS), Interrupted aortic arch (IAA), Ebstein anomaly |
| 4 | “Obstructive lesions of valves and vessels” | Pulmonary stenosis, Aortic stenosis, Coarctation of the aorta, Bicuspid aortic valve, Pulmonary artery branch stenosis |
| 5 | “Simple defects of septation and patent arterial duct “ | Patent ductus arteriosus (PDA), VSD, ASD. |
| 6 | “Other great vessel and coronary artery lesions” | Aortic arch anomalies, Coronary artery anomalies, Partial anomalous pulmonary venous return, Pulmonary artery sling |

**Table 2.** Severity categorization.

| Category | Defects included |
| --- | --- |
| Severity I (SI) | Single ventricle pathway/physiology, Hypoplastic left heart, Hypoplastic right heart, Ebstein anomaly, Tricuspid atresia, Mitral atresia. |
| Severity II (SII) | Pulmonary valve atresia, Common arterial truncus, Atrioventricular septal defects (common AV canal), Aortic valve atresia/stenosis, Transposition of the great arteries, Tetralogy of Fallot (including severe type with pulmonary atresia & major aortopulmonary collateral arteries (MAPCA)), Total anomalous pulmonary venous return, Coarctation of the aorta, Double outlet right or left ventricle, Discordant atrioventricular connection, Ivemark atrial isomerism, Other abnormal cardiac connections, Aortopulmonary window, Subaortic valve stenosis, Malformations of coronary arteries, Atresia of aorta, Interrupted aortic arch, Supravalvular aortic stenosis, Abnormal pulmonary venous connections<br>Without additional CHD subgroups classified as severity I. |
| Severity III (SIII) | Ventricular septal defect, Atrial septal defect, Pulmonary valve stenosis, Mitral regurgitation, Mitral stenosis, Arterial tortuosity syndrome<br>Without additional CHD subgroups classified as severity I or II. |

### Statistical Analysis

Data was analyzed using SPSS ® statistical software *(Version 27*.*0, International Business Machine Corporation ®, 2020)* and Microsoft Office Excel *(Version 16*.*69*.*1, Microsoft © 2022)*. Categorical variables were described with frequencies and percentages, and nominal variables (age) was presented with median and range. Statistical analysis was performed using Chi-Square tests and Fisher’s Exact tests when analyzing 2 categorical variables. Significance values were derived from the exact significant Pearson Chi-Square test value, and when cell counts were less than 5, then value from the Fisher’s Exact test was used. For all tests, significance was set at P<0.05.

The Institution Review Boards at Sidra Medicine and Weill Cornell Medicine – Qatar approved the study. Informed consent was not required because data were collected retrospectively from the registry database.

## Results

At the time of data collection for this study, the research registry had data for 85 consented patients with CHD, channelopathies and cardiomyopathies collected between July 2019 and October 2022. It should be noted that the registry does not include all CHD patients attended at Sidra Medicine during the mentioned duration. From the 85 patients, we excluded 9 subjects for the following reasons: 6 had pure cardiomyopathies and 3 had channelopathies. The final sample size used in the analysis was 76 subjects, which included 43 males (56.6%) and 33 females (43.4%). Table 3, 4 and 5 describes the demographics, defect types, and classification of CHD cases according to severity and anatomic categories. Of the 76 subjects, 64 are currently living with a median age of 3.53 years, and 12 are deceased, with a median age at death of 10 months. Most of the subjects were from the MENA region (excluding Qatar) (52.6%), followed by Southeast Asian (30.3%) and Qatari (14.5%). The three most common CHD types were VSD, ASD, and TGA. About 25% of the subjects were categorized into Severity I (high perinatal mortality), 64.5% to Severity II, and 10.5% to Severity III (low perinatal mortality) category.

**Table 3:** Subjects’ living status and age.

|  | N or median | % or Range |
| --- | --- | --- |
| **Living status & age** |  |  |
| Living | 64 | 84.2% |
| Current Age (Living Patients) | 3.53 years | 0.41-18.26 years |
| Deceased | 12 | 15.8% |
| Age at Death (Deceased Patients) | 0.83 years (10 months) | 0.17-16 years |

**Table 4:** Subjects’ demographics and CHD distribution.

| Gender | N | % |
| --- | --- | --- |
| Male | 43 | 56.6% |
| Female | 33 | 43.4% |
| **Nationality** |  |  |
| Qatari | 11 | 14.5% |
| Middle Eastern/North African (Non-Qatari) | 40 | 52.6% |
| South Asian | 23 | 30.3% |
| Other | 2 | 2.6% |
| **Ten most commonly presenting CHD types (Patients may present with >1 CHD, thus total % may exceed 100)** |  |  |
| Ventricular Septal Defect | 26 | 34.2% |
| Atrial Septal Defect | 24 | 31.6% |
| Transposition of the Great Arteries | 17 [D-TGA: N=15; L-TGA: N=2] | 22.4% |
| Patent Ductus Arteriosus | 17 | 22.4% |
| Pulmonary Stenosis | 17 | 22.4% |
| Tetralogy of Fallot | 14 | 18.4% |
| Atrioventricular Septal Defect (AVC) | 14 | 18.4% |
| Pulmonary Atresia | 11 | 14.5% |
| Double Outlet Right Ventricle | 9 | 11.8% |
| Right Ventricular Hypoplasia | 8 | 10.5% |

**Table 5:** CHD severity and anatomical categorization.

| **CHD Severity categorization** |  |  |
| --- | --- | --- |
| SI | 19 | 25% |
| SII | 49 | 64.5% |
| SIII | 8 | 10.5% |
| **CHD anatomical categorization** (Patients may be in >1 group, thus total % may exceed 100) |  |  |
| 1 | 10 | 13.2% |
| 2 | 50 | 65.6% |
| 3 | 26 | 34.2% |
| 4 | 21 | 27.6% |
| 5 | 6 | 7.9% |
| 6 | 1 | 1.3% |

### Consanguinity and CHD

Of the 76 subjects, 33 (44.3%) were born from consanguineous marriage. The distribution of the degree of parent’s relationship among subjects born from consanguineous marriages showed that 24 (31.58%) subjects were born by parents who were first-degree cousins, while second- and third-degree parental cousins were reported in 7 (9.2%) and 1 (1.3%) of the subjects respectively (Table 6 and Figure 1). There was a significant association between first degree cousins and congenital heart defects (P<0.05). Our results also showed that a higher proportion of consanguinity was found among non-Qatari Arabs from the MENA region. However, this association was not statistically significant (p>0.05).

**Table 6:**
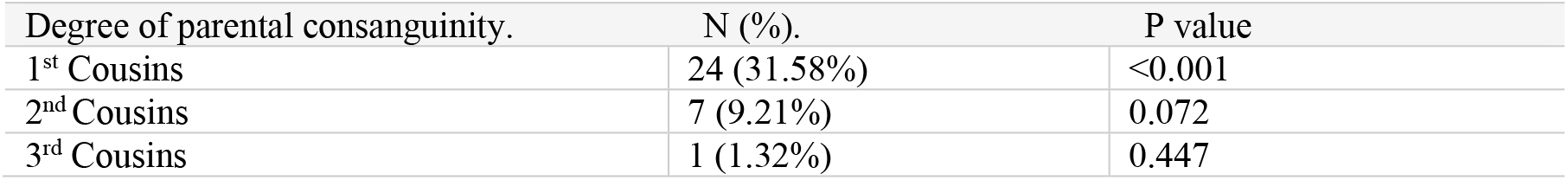
Degree of parental consanguinity and CHD.

| Degree of parental consanguinity. | N (%). | P value |
| --- | --- | --- |
| 1<sup>st</sup> Cousins | 24 (31.58%) | $< 0.001$ |
| 2<sup>nd</sup> Cousins | 7 (9.21%) | 0.072 |
| 3<sup>rd</sup> Cousins | 1 (1.32%) | 0.447 |

**Figure 1:**
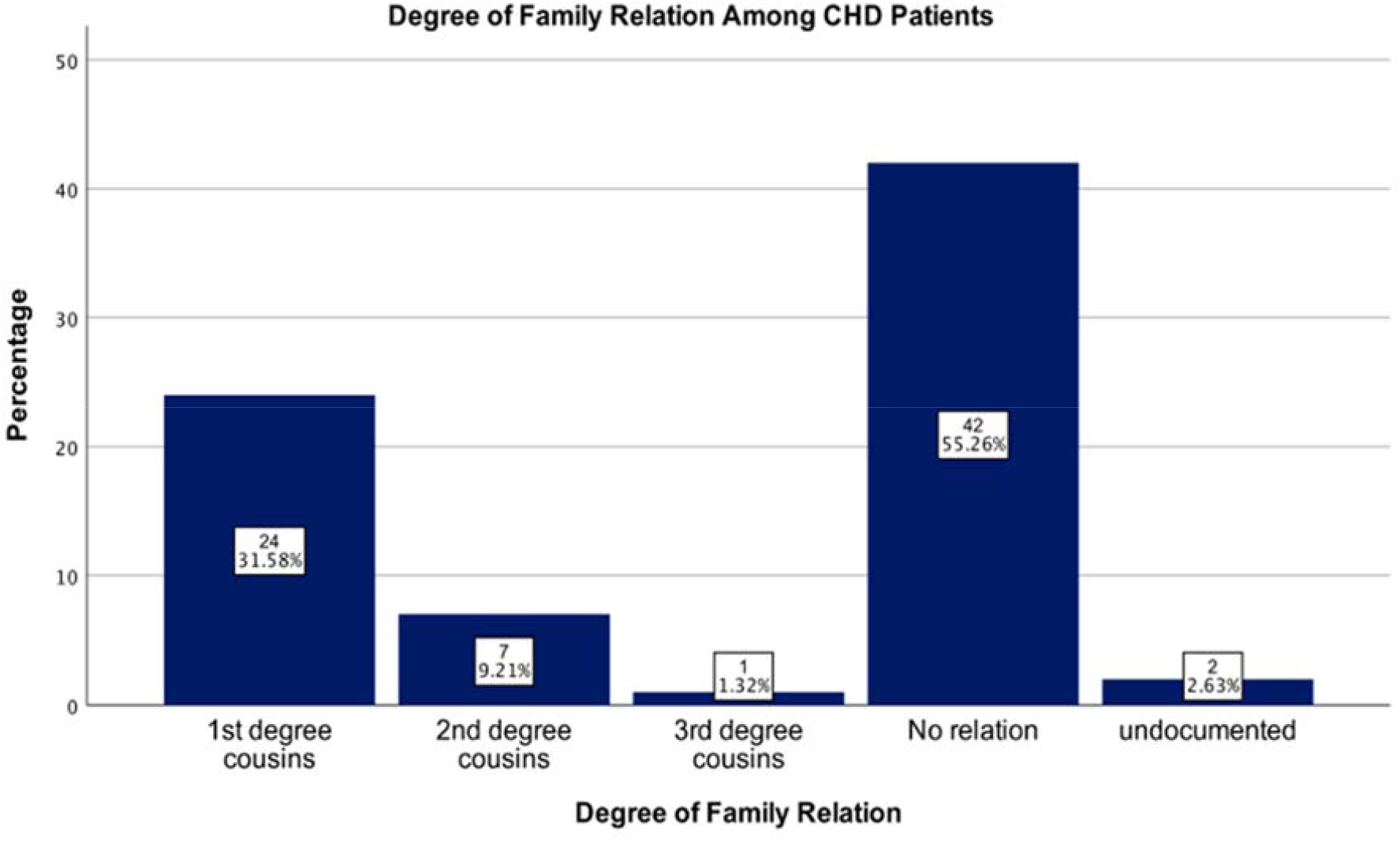
Parental consanguinity among patients with CHD.

The frequency of Individual CHD, and their association with parental consanguinity was also investigated. The highest proportion of parental consanguinity (30.3%) was found among patients with VSD. However, this association was not statistically significant (Table 6). There was a significant association between parental consanguinity and patients with PDA (p value <0.05).

### Non-genetic Risk Factors and CHD

We also explored the association between of some of the non-genetic risk factors like maternal diabetes mellitus and maternal age with CHD. Only 58 subjects with adequate maternal history were included in this analysis. About 38% subjects in this cohort were born to mothers who had Gestational Diabetes Mellitus, while 6.9% to mothers with Pre-Gestational Diabetes Mellitus (Table 7). About 68.2% of subjects born to mothers with gestational DM, were classified into severity II CHD category while all the patients born to mothers with pre-gestational DM were classified into severity category II CHDs. There was no significant statistical association between severity of CHD and maternal diabetes mellitus. We also explored the association between maternal diabetes (both pre-gestational and gestational) and CHD types. Only DORV was significantly associated with maternal Diabetes Mellitus (p< 0.05). Given the small number of patients under gestational and pregestational Diabetes Mellitus, further analysis to determine association between type of maternal diabetes and individual CHD lesions was not conducted.

**Table 7:** Maternal diabetes mellitus and CHD.

| *Severity (SI-III)* | *Maternal Diabetes #* | *Pre-gestational diabetes #* | *Gestational diabetes #* | *P value* |
| --- | --- | --- | --- | --- |
| Severity I | 5 | 0 | 5 | 0.545 |
| Severity II | 19 | 4 | 15 | 0.580 |
| Severity III | 2 | 0 | 2 | 0.675 |
| Total | 26 | 4 | 22 | 0.653 |

The age of majority (53.4%) of mothers of patients with CHDs were between 26 and 35 years, while 24.1% were aged above 36 years of age. There was no significant association between severity of CHD and maternal age. None of the mothers of the patients with CHDs smoked cigarettes or drunk alcohol.

### Outcomes of CHD

The documented 6-month outcomes were as follows: 81.5% of subjects had history of admission to the neonatal intensive care unit, 68.9% underwent surgical intervention, 53.5% underwent interventional catheterization, 61.8% were hospitalized (not including NICU or elective admissions), and 4.1% died (Table 8). Of these outcomes, only interventional catheterization was associated with category 3 anatomic classification [Severe obstructions (hypoplasias and atresias)] in the first six months of life (p< 0.05). The most common reason for hospitalization was infection, documented in 76.2% of subjects. The most common types of infection included bronchiolitis, urinary tract infections and pneumonia.

**Table 8:** 6-Month Outcomes of patients with CHD.

| **Outcome** | **N** | **%** |
| --- | --- | --- |
| NICU | 44 | 81.5% |
| Surgery | 51 | 68.9% |
| Interventional catheterization | 38 | 53.5% |
| Hospitalization | 42 | 61.8% |
| Death | 3 | 4.1 |

### Comorbidities Among Subjects with CHD

This study also looked at the comorbidities affecting subjects with CHDs. The findings shows that the leading comorbidities were those of cardiac origin followed by those affecting the renal, gastrointestinal, and endocrine systems (Figure 2). Failure to thrive, sepsis, and arrhythmia represented the highest individual acquired comorbidities among subjects with CHD at 58.1%, 27.0%, and 25.7%, respectively. On the other hand, gastric anomaly, immunodeficiency, and renal anomalies were the leading congenital comorbidities among subjects with CHD, at 18.9%, 14.9%, and 10.8 %, respectively. Specifically, patients with TOF showed the highest rates of comorbidities, followed by PDA and anomaly of the pulmonary artery. When comparing the relative risks for having individual comorbid conditions among female patients with CHD versus male patients with CHD, female patients are at higher risk of having hypertension, pulmonary hypoplasia, immunodeficiency, anemia, and abnormality (RR >1.5).

**Figure 2:**
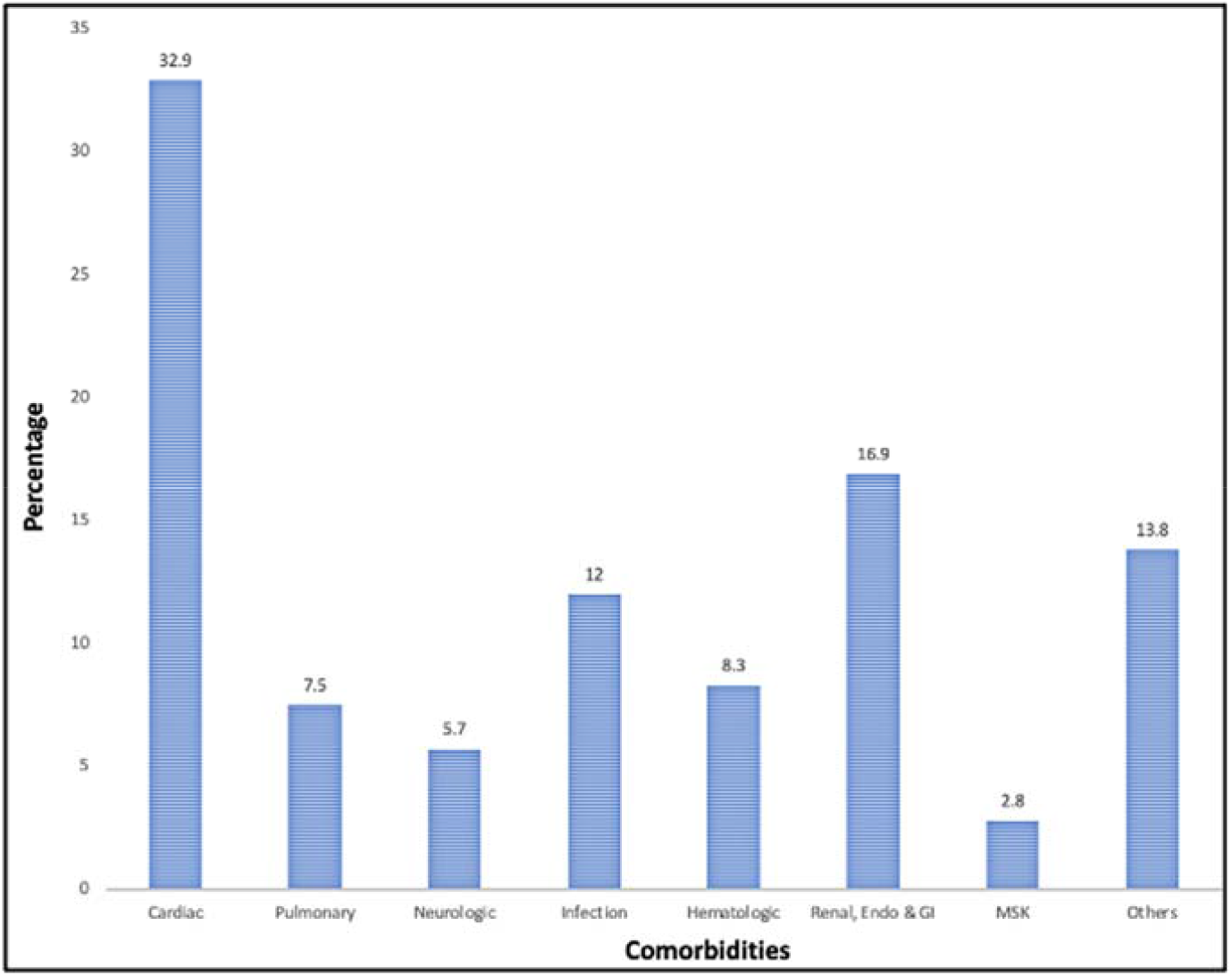
Comorbidities and CHD.

## Discussion

This study provides a preliminary analysis on the epidemiology of CHD in Qatar. Our results shows that majority of the patients are from the MENA region followed by patients from South Asia.

The most common CHD subtypes in our study was VSD followed by ASD, TGA and PDA. This pattern is comparable with findings from other studies.^2,18^

This study also looked at the rate of parental consanguinity among subjects with congenital heart defects. Our findings revealed that the MENA region had the highest rate of CHD patients born from consanguineous marriages. Of the 44.3% subjects born by consanguineous parents, 31.58% were born by first-degree cousins. The association between subjects born by first-degree parental cousins and CHD, was significant (P<0.05). Parental consanguinity was highest among subjects with VSD, followed by ASD. Consanguinity rates in the middle eastern countries ranges between 20 and 50%.^19^ Previous studies have shown that these countries have the highest rate of consanguinity among patients with CHDs.^2,20^ This association is especially higher among first-degree parental cousins.^21^ Consanguineous marriages pose a significant genetic risk for the offspring because of the inherited identical chromosomal segments from both parents.^22^ First-degree cousins’ marriage increases the risk for cardiac malformation to 5-8%.^23,24^ Several genes have been implicated for CHD, but incomplete penetrance and variable expression of the phenotypes complicates the association between the genetic mutations and CHDs.^25^ Recessive gene mutations may play a role in the causation of CHD in first-cousin marriages, even though the exact mechanism is not fully understood.^26^

Maternal diabetes mellitus is considered as one of the most important risk factors for CHD. Of the CHD patients born to diabetic mothers in this study, majority were born by mothers who had gestational diabetes mellitus and only a small number were born by mothers who had pre-gestational diabetes mellitus. VSD, ASD and PDA were the most common defects seen in infants of diabetic mothers. There was no significant association between maternal diabetes (pre-gestational or gestational) and CHD. Because of the small sample size, further analysis on the association between individual type of maternal diabetes mellitus (pre-gestational and/or gestational) and CHD was not conducted. Previous studies have shown that VSD, ASD and ToF are associated with pre-gestational DM.^27,28^ Both pre-gestational and gestational diabetes mellitus are detrimental for the fetus.^29^ The first 6-8 weeks of human developmental are extremely crucial for the cardiac development, and elevated levels of HbA1c or fasting serum glucose during this time may be associated with a higher likelihood of CHD in the neonate.^11^ Women with pre-existing diabetes mellitus, therefore, are at a slightly higher risk of giving birth to newborns with CHD than those with gestational diabetes.^11,30^ Some of the experimental-supported mechanisms by which maternal diabetes mellitus alters cardiac development include glucose-mediated disturbances of left-right patterning, increased apoptosis due to oxidative or other cellular stress, nitric oxide signaling deficiencies, and alterations of neural crest cell formation and migration.^31,32,33,34^

Older maternal age, particularly above the age of 35 years, has been linked to several birth defects and chromosomal abnormalities in infants. We found no association between CHD and maternal age. Our findings are consistent with a large-scale study conducted in England which concluded that, there was no evidence of increased risk of CHD in women aged 35 years and above.^35^

Of the 74 subjects analyzed for six-month outcomes, only 54 had information on NICU status after birth. Among these patients, majority (81.5%) had a history of being admitted to the NICU. The majority of patients had multiple CHD lesions, reflecting their complexities. It is well known that children with complex congenital heart defects require highly specialized, resource-intensive care facilities, including NICU.

Our results also shows that 68.9% of the subjects in this study underwent surgical intervention, and 53.5% underwent interventional catheterization within the first six-months of life. Interventional catheterization was more likely to be performed in patients with Embryological Category 3 (severe obstructions, hypoplasias, atresias). Current trends on the choice of interventions in patients with CHD show an increase in surgical intervention and earlier primary correction for severe defects, while catheter-based interventions are preferred for simpler defects.^14^

About 62% of the subjects in this study had history of hospitalization by the age of six months. Hospitalization was not found to be significantly associated with any variable like nationality, anatomical or severity categorization. The most common causes of hospitalization included infection and sepsis, followed by heart failure, and hemodynamic compromise. Bronchiolitis due to RSV, rhinovirus, and parainfluenza were the most common infections. Elsewhere, pleural effusion, pericardial effusion, arrhythmia, pneumonia, and bronchiolitis have been reported as the common causes of hospitalization for patients with CHD.^36^ Current guideline recommendations indicate that RSV prophylaxis should be given to all children aged ≤12 months .^37^

Neurologic complications were also noted in our sample, with both stroke and seizures documented in 9.5% of the subjects. Beginning in utero, abnormal circulatory patterns due to CHDs predispose a fetus to poor delivery of oxygen rich blood to the brain, negatively affecting fetal brain growth. Stroke and focal brain injury, likely due to abnormal brain development and impaired cerebral circulation, may further contribute to neurodevelopmental deficit which is a known complication in children with CHD. Seizures are common acquired brain injury symptom in patients with CHDs, and can occur post-operatively, and sometimes secondary to acute hemorrhagic or ischemic stroke.^38^ Stroke has been reported as a common neuroimaging finding, and the most common pre-operative injury among patients with CHDs, particularly in D-TGA patients.^38^

About 16% of subjects in the cardiac registry are deceased, with about 4% of the deaths occurring in the first six months of life. The cause of death could not be obtained. The mortality was not associated with nationality, parental consanguinity, severity scale categorization, or embryological categorization. The overall death rate in Qatar appears to be comparable to that found in a 15-year study done in Norway, for which 11.7% of patients died 3-18 years post follow-up.^13^ In the Norwegian study, 52.4% of deaths occurred during the neonatal period, whereas none of the deaths occurred in the first month of life amongst the Qatar CHD population.

Almost every subject in this study had other disease in addition to the underlying CHD lesion, with TOF carrying the heaviest burden of having the highest rate of comorbidities. Generally, arrhythmias and failure to thrive were the most common cardiovascular comorbidities among subjects included in our study. The common comorbidities in Patients with TOF included HTN, arrhythmia, stroke, and anemia. The anatomy of ToF allows the mixing of deoxygenated blood with a subsequent right-to-left shunting in the systemic circulation. With less blood flow going into the lungs, hypoxia is an inevitable outcome, which drives the pathological change and ultimately leads to the common complications seen in these patients. The overall estimates of the comorbidities were lower than most of the previous studies.^39,40^ However, most of these studies had large sample sizes and consisted of an older cohort of patients which most likely contributed to the difference seen in our study. It is well known that as we age, our physiological processes slow down, resulting in global dysfunction in body systems and the consequent emerging comorbidities.^41^

Findings from this study should be interpreted considering several limitations. Firstly, it is a preliminary study of a newly developed cardiac registry, with a small sample size of CHD cases so far. As a result of the small sample size, several variables had fewer number of patients further limiting statistical analysis. A second limitation is the inconsistency in the documentation on patient charts due to patients seeking care across several hospitals which limited analyzing all the possible variables in this study.

## Conclusions

In conclusion, this retrospective study adds to the literature information on the epidemiology of congenital of heart diseases in Qatar. As in other studies, the most common CHD type in this study was VSD followed by ASD and TGA. The rate of parental consanguinity was generally high among patients with CHDs. The pattern of non-genetic risk factors, comorbidities, and outcomes in patients with CHD was like those previously reported in other studies.

## Data Availability

The authors confirm that the data supporting the findings of this study are available within the article

## Nonstandard Abbreviations and Acronyms

MAPCA: Major aortopulmonary collateral arteries
MENA: Middle East and North Africa
DILV: Double inlet left ventricle
DORV: Double outlet right ventricle
TAPVR: Total anomalous pulmonary venous return
AVSD: Atrioventricular septal defect
HLHS: Hypoplastic left heart syndrome
IAA: Interrupted aortic arch

